# Differential COVID-19 mortality in the United States: Patterns, causes and policy implications

**DOI:** 10.1101/2023.01.09.23284358

**Authors:** Michael A. Stoto, Samantha Schlageter, Duccio Gamannossi degl’Innocenti, Fabiana Zollo, John D. Kraemer

**Affiliations:** Department of Health Management and Policy, Georgetown University, Washington DC, USA; Università Cattolica del Sacro Cuore, Milan, Italy; Ca’ Foscari University of Venice, Venice, Italy

## Abstract

A “two Americas” narrative emerged in the summer of 2021: one with high demand for COVID-19 vaccines, and a second with widespread vaccine hesitancy and opposition to mask mandates. But our analysis of excess mortality shows that the U.S. has been a divided nation at least since the start of the pandemic. Through April, 2022, there were 1,335,292 excess deaths associated with COVID-19, 37% more than reported as such. After the first wave, death rates in the South were more than double those in the Northeast; 45% of deaths were in the South, with 38% of the population.

While some regard vaccination and other measures as matters of personal choice, the population impact is striking. If every region had the same mortality rate as the lowest regional rate in each period, more than 418,763 COVID-19 deaths were “avoidable,” more than half (58%) in the South and almost half before vaccines were available. These results show that population-based COVID-19 policies can still play an important role in protecting those most vulnerable to severe disease and death and reducing the spread of the virus.

This example illustrates the importance of excess mortality measures as part of a comprehensive surveillance system. Official mortality counts rely on complete recording of COVID-19 as a cause of death, but COVID-19 deaths are under reported for many reasons. Indeed, the proportion of COVID-19 deaths reported as such varied markedly over time, and from 67% in the West to 87% the Northeast. In 2022, some regions cut back on testing making it harder to see a re-emergence of COVID-19 in those places. More extensive surveillance based on wastewater testing and other means that do not depend on testing are needed to get a more accurate picture. Excess mortality estimates are more tenuous years beyond the pre-pandemic period.

## Introduction

As analysts seek to understand why COVID-19 mortality in the United States lags behind other countries (1), the focus has been on differentials in vaccine uptake (2). A narrative of “two Americas” emerged in the summer of 2021. Some states, especially those that leaned towards the Democratic party (3), demonstrated a high demand for the COVID-19 vaccine. Other states resisted vaccines and later opposed mask and vaccine mandates. The second America was concentrated in Southern states and rural areas, especially those that voted for Donald Trump. Since then, cases, hospitalizations, and deaths increased dramatically in the second America (4). This narrative not only shapes how we understand what happened and why, but also influences analyses of what should, or could, be done to control future outbreaks.

In an earlier paper, we found that reality is more complex. Stark regional differences in COVID-19 mortality existed from the start of the pandemic, both in cases and deaths as well as testing and vaccine uptake (5). In this paper we extend that analysis into the early months of 2022 and incorporate new regional data on vaccine uptake and mask use. The extended analysis demonstrates regional patterns in these variables as well in mask use and vaccine uptake throughout the pandemic.

Reported cases, hospitalizations, and deaths are known to be substantial underestimates depending on test availability, patient and physician awareness and attitudes, hospital resources, and other factors (6,7). COVID-19 awareness and concern (4,8,9), as well as test availability and use (10–13), vary markedly during the pandemic and throughout the U.S. We avoid these problems by focusing on “excess mortality” estimates, which are based on the difference between total number of deaths experienced in a given week and the number that would have been expected based on earlier time periods. These methods have been used in the U.S. (14–17), Italy (18), Canada (19) and many other countries. Both the World Health Organization (1) and the Institute for Health Metrics and Evaluation (20) have used the excess mortality approach to examine the impact of COVID-19 around the globe. This analysis illustrates the strengths and weaknesses of excess mortality estimates, and considers their role as part of the public health surveillance toolbox.

## Materials and Methods

As in our original paper (5), this analysis is based on state-level weekly excess mortality calculations published by CDC (21). Farrington surveillance algorithms, which use over-dispersed Poisson generalized linear models with spline terms to model trends in counts, accounting for seasonality, were implemented for each jurisdiction (states, plus the District of Columbia and New York City). Further details about these methods have been published by the CDC (13). Logically, the expected number of deaths should be based on data before the pandemic, but CDC’s most recent excess mortality estimates include part of the pandemic period in the baseline. Consequently, this analysis is based on the expected mortality forecasts that CDC published in September 2021, the same dataset we used in our previous paper (5). In particular, deaths from the CDC dataset published in September 2021 were used to calculate expected deaths up until September 25, 2021. After this date, we calculated the relative change in expected deaths in each state between September 2019 and April 2020 and applied this to the September 2021 to April 2022 timeframe.

For the sake of presentation, we have divided the study period into six periods based on overall U.S. COVID-19 cases and mortality. These periods correspond roughly to the initial phase of the pandemic, the summer of 2020, the Alpha wave, the Spring of 2021, the Delta wave, and the Omicron wave, and we use these terms as a shorthand to describe the phases.

In order to visualize patterns in COVID-19 mortality and risk factors, the results are presented in the four standard U.S. Census regions (see map in supplemental material). Other *ad hoc* groupings could exaggerate or minimize differences. The states in the Northeast and South are relatively similar, but there is more internal variation in the West, where California is an outlier in some periods. The calculations were done by week and state and aggregated by adding the estimated of excess deaths in each region and time period.

“Avoidable mortality” was calculated by assuming that the lowest excess mortality rate experienced by any region in each period could have been achieved by all regions. Because the first period was unique we did not include the first period in these calculations, but as a sensitivity analysis we estimated the numbers of deaths that could have been avoided in Period 1 as one-half of the difference between the actual rates and those in the West, the region with the lowest excess mortality rate in that period.

Vaccination data are calculated by the authors for each of the Census regions based on the CDC Data Tracker (12). In particular, we calculate (1) the proportion of the population aged 65 and older who are fully vaccinated (i.e. have received two doses of the Pfizer-BioNTech or Moderna vaccines or 1 dose of the Johnson & Johnson/Janssen vaccine), (2) the proportion of the population aged 18 to 64who are fully vaccinated, and (3) the proportion of the fully vaccinated population aged 65 and older who have received at least one booster dose. Because vaccination is by its nature cumulative, we present cumulative proportions on three dates: June 26, 2021; December 4, 2021; and April 30, 2022, the ends of period 4, 5, and 6 respectively.

Behavior data for mask use are calculated by the authors using the COVID-19 Trends and Impact Survey (CTIS) (22). The survey was administered by the Delphi Group at Carnegie Mellon University in partnership with Facebook and deployed in 13 waves from April 6, 2020, to June 25, 2022 (see details in supplemental material). The survey participants were sampled from Facebook users. To obtain a representative sample of the United States population, Facebook used its own demographic data to calculate statistical weights for each survey participant (23). As of November 2021, the average daily number of survey responses was about 40,000, and the total number of responses was over 25 million. The survey asked participants questions about COVID-like symptoms, vaccine acceptance, their behavior (e.g., social distancing, mask use), mental health, and economic and health impacts of the pandemic on their life. Here we calculate for each of the Census regions the weekly proportion of individuals wearing a mask from September 8, 2020 (Period 2) to June 25, 2022 (Period 6). We do so by combining questions C14 (“In the past 5 days, how often did you wear a mask when in public?” in Waves 4-7), and C14a (“In the past 7 days, how often did you wear a mask when in public?” in Waves 8-13).

## Results

Between Jan. 3, 2020 and April 30, 2022, public health officials reported 978,567 COVID-19 deaths in the U.S. According to our calculations, however, the excess mortality associated with COVID-19 totaled 1,335,292, during that period, 37% more COVID-19 deaths than reported. As can be seen in Figure 1, there are clear differences among the regions over time, especially between the Northeast and South.

**Figure 1.**
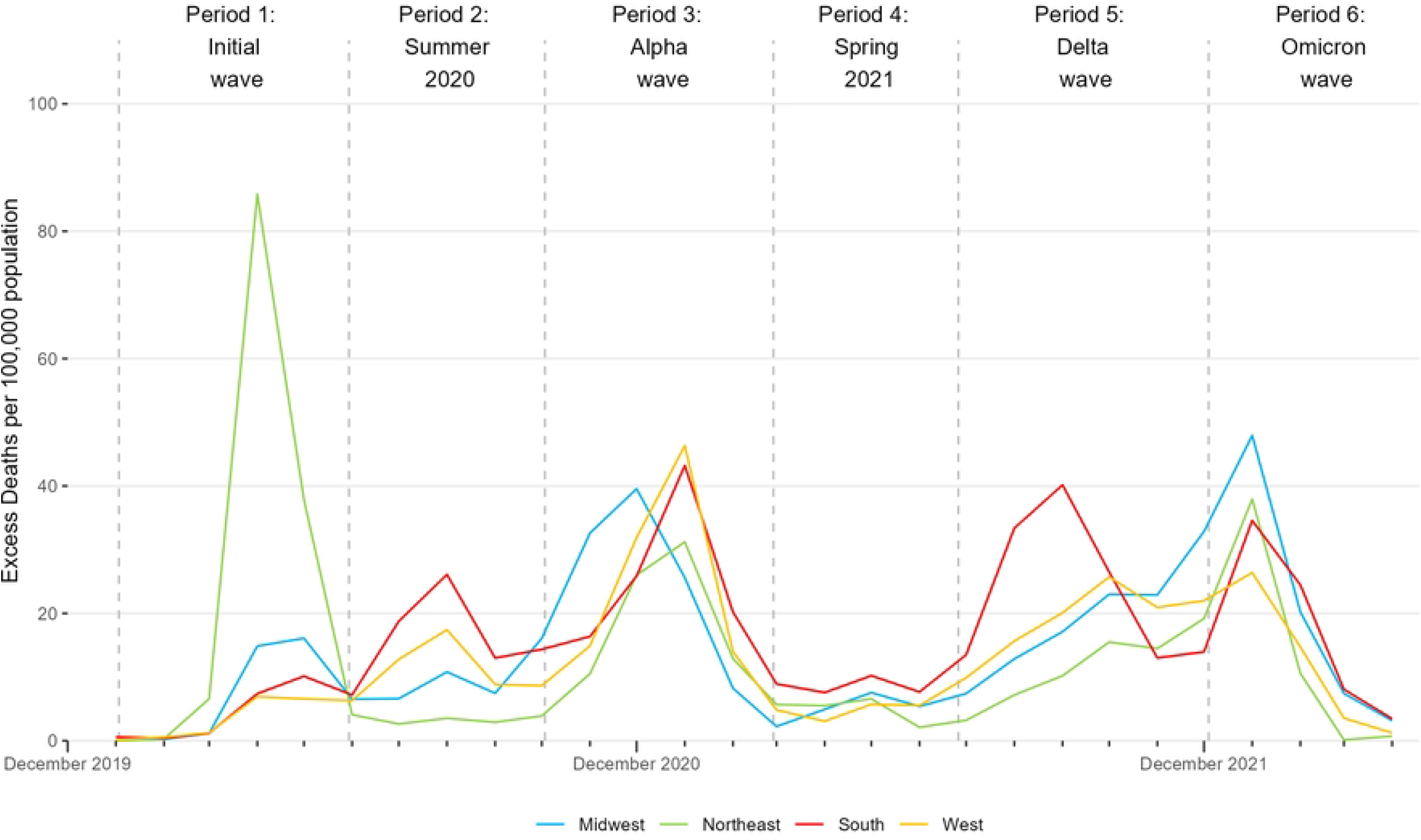
Excess mortality per 100,000 population by week and region, U.S., January 3, 2020 – April 30, 2022. The periods are denoted by vertical lines corresponding to May 30, 2020, Oct. 3, 2020, Feb. 27, 2021, June 26, 2021, and Dec. 4, 2021. Source: authors’ calculations based on CDC data.

In the initial wave of the pandemic, before May 30, 2020, deaths in the Northeast dominated the other regions; subsequently, deaths in this region were lower than in the rest of the United States. Deaths were relatively high in the South during the summer of 2020 (May 31 – Oct. 3) and the Delta wave (June 26 – Dec. 4, 2021). The Midwest experienced relatively large numbers of deaths in the first half of the Alpha wave (Oct. 4, 2020 – Feb. 27, 2021) and the Omicron wave (Dec. 4, 2021– Apr. 30, 2022).

Figure 2 shows that the majority of these deaths were during the Alpha (28%), Delta (25%), and Omicron (19%) waves. In the initial wave (before May 31, 2020), approximately 56% of deaths were in the Northeast; subsequently, 45% were in the South.

**Fig 2.**
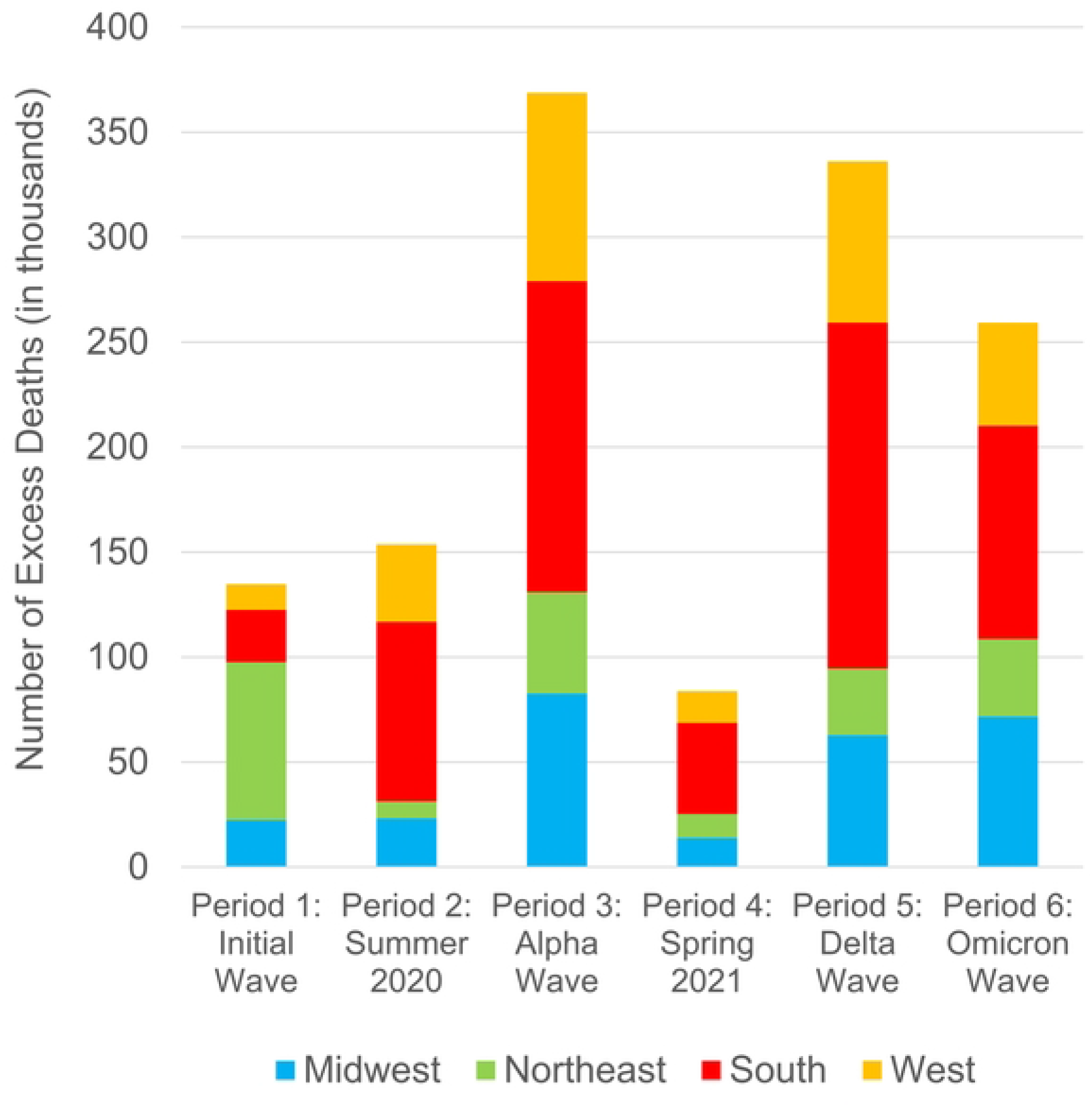
Number of excess deaths by period and region, U.S., January 3, 2020 – April 30, 2022. The height of each bar represents the total U.S. excess deaths in each period and the colored sections represent the number of excess deaths in each region during the period. Periods are defined as follows: Period 1 (January 3 to May 30, 2020), Period 2 (May 31 – October 3, 2020), Period 3 (October 4, 2020 to February 27, 2021), Period 4 (February 28 to June 26, 2021), Period 5 (June 27 to December 4, 2021), and Period 6 (December 5, 2021 to April 30, 2022). Source: authors’ calculations based on CDC data.

Table 1 displays excess mortality rates per 100,000 population per day by region and period. Before May 31, 2020, the daily mortality rate in the Northeast (0.888 per 100,000) was 3.2 times the national rate while the rate for the South (0.134 per 100,000) was 0.49 times the national rate. Subsequently, the South experienced COVID-19 mortality 3% to 46% higher than the national rate, whereas the Northeast’s rate was 11% to 70% lower.

**Table 1.** Excess and avoidable mortality by region and time period, U.S., January 3, 2020 – April 30, 2022.

|  | *Period 1*<br>*(1/3–*<br>*5/30/20)* | *Period 2*<br>*(5/31–*<br>*10/3/20)* | *Period 3*<br>*(10/4/20–*<br>*2/27/21)* | *Period 4*<br>*(2/28–*<br>*6/26/21)* | *Period 5*<br>*(6/27–*<br>*12/4/21)* | *Period 6*<br>*(12/5/21–*<br>*4/30/22)* | *Entire*<br>*period* |
| --- | --- | --- | --- | --- | --- | --- | --- |
| *Excess mortality by region and period* |  |  |  |  |  |  |  |
| Midwest | 22,367 | 23,212 | 82,676 | 13,883 | 62,749 | 71,674 | 276,561 |
| Northeast | 75,216 | 8,029 | 48,243 | 11,457 | 31,785 | 36,848 | 211,578 |
| South | 24,886 | 85,453 | 148,191 | 43,410 | 164,827 | 101,655 | 568,422 |
| West | 12,206 | 36,986 | 89,608 | 15,064 | 76,727 | 49,141 | 279,732 |
| U.S. | 134,675 | 153,680 | 368,718 | 83,814 | 336,088 | 259,317 | 1,336,292 |
| *Excess mortality per 100,000 population per day by region and period* |  |  |  |  |  |  |  |
| Midwest | 0.221 | 0.267 | 0.815 | 0.169 | 0.565 | 0.707 | 0.473 |
| Northeast | 0.888 | 0.111 | 0.570 | 0.167 | 0.343 | 0.435 | 0.434 |
| South | 0.134 | 0.537 | 0.798 | 0.289 | 0.811 | 0.548 | 0.531 |
| West | 0.106 | 0.374 | 0.776 | 0.161 | 0.606 | 0.425 | 0.420 |
| U.S. | 0.276 | 0.368 | 0.757 | 0.212 | 0.630 | 0.532 | 0.476 |
| *Excess mortality as a proportion of reported COVID-19 deaths* |  |  |  |  |  |  |  |
| Midwest | 85% | 61% | 93% | 98% | 60% | 65% | 75% |
| Northeast | 83% | 101% | 105% | 127% | 56% | 83% | 87% |
| South | 79% | 67% | 84% | 62% | 66% | 61% | 70% |
| West | 76% | 57% | 84% | 67% | 52% | 62% | 67% |
| U.S. | 82% | 66% | 89% | 78% | 61% | 66% | 73% |
| *Avoidable excess mortality by region and period* |  |  |  |  |  |  |  |
| Midwest | - | 13,597 | 24,906 | 164 | 24,687 | 27,550 | 90,904 |
| Northeast | - | 0 | 0 | 414 | 0 | 825 | 0 |
| South | - | 67,855 | 42,453 | 18,299 | 95,162 | 20,893 | 244,663 |
| West | - | 26,033 | 23,796 | 0 | 33,367 | 0 | 83,196 |
| U.S. | - | 107,486 | 91,156 | 18,877 | 153,215 | 49,268 | 418,763 |

Table 1 also shows how the ratio of estimated to reported COVID-19 deaths varies among the regions over time. The West (67%) and South (70%) report the lowest fraction. In Period 1 the proportions were similar in all regions (approximately 82%), but they vary markedly afterward, in particular dropping to less than 60% in the West in some periods. The overall proportion is highest in the Northeast (87%). The proportion is greater than 100% in some periods, probably because some people who died of another primary cause had a positive COVID-19 test and were included in the reported counts (14), as called for by the National Center for Health Statistics (24). A recent analysis attributed the more accurate coding of COVID-19 deaths in the New England states (which are in the Northeast region) to well-run and funded public health departments, excellent hospitals, and state medical examiners who ensure death certificate information is both accurate and timely (25). The report also noted that some states in the region had large increases in deaths from overdoses rather than COVID-19, indicating a weakness of the excess mortality approach.

Because the rates of excess mortality vary so markedly, one can calculate how many deaths could have been avoided if each region had the same rates as the lowest region in each time period, a standard demographic calculation. We assume that no deaths were avoidable in the first period, before much was known about treating or preventing COVID-19. Subsequently, we counted as “avoidable” the difference between the actual rates and the lowest regional rate in each period. This calculation indicates that 418,763 COVID-19 deaths were avoidable. As can be seen in Figure 3, more than half of these avoidable deaths (58%) were in the South. Almost half (47%) occurred during the Alpha wave and the summer before it, that is between May 31, 2020 and February, 2021. A similar number (48%) occurred during the Delta and Omicron waves, between June 27, 2021 and April 30, 2022.

**Fig 3.**
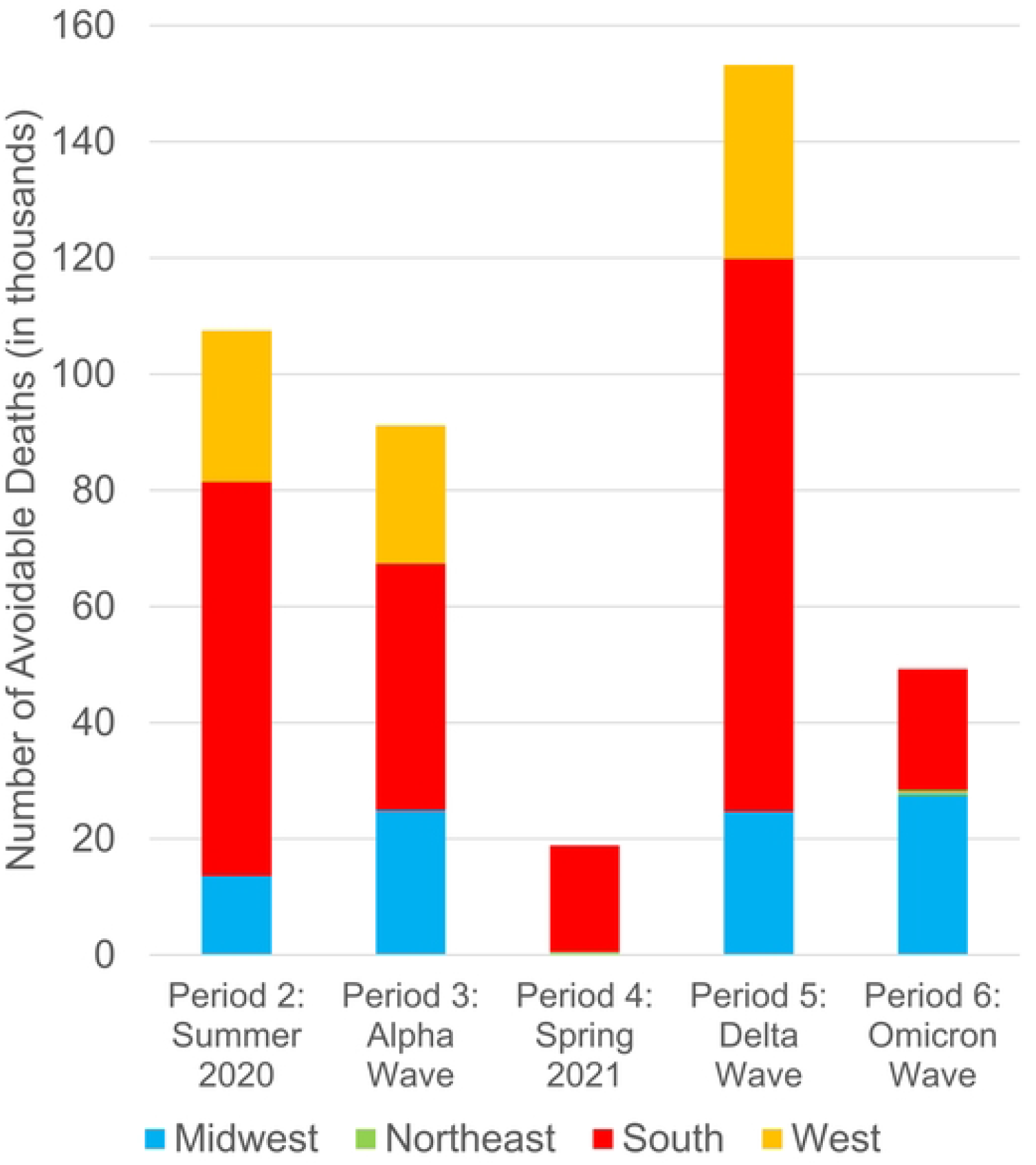
Number of avoidable deaths by period and region, U.S., January 3, 2020 – April 30, 2022. The height of each bar represents the total U.S. avoidable deaths in each period and the colored sections represent the number of avoidable deaths in each region during the period. Periods defined as in Figure 2. Source: authors’ calculations based on CDC data.

The initial wave was excluded from the avoidable mortality calculations because one might argue that too little was known at this time about how to prevent transmission. As a sensitivity analysis, we calculated the number of avoidable deaths in Period 1 as one-half of the difference between the actual rates and those in region with lowest excess mortality rate in that period. With this assumption the proportion of avoidable deaths in the South was still high, 54%, and the total avoidable deaths increases to 461,600.

## Discussion

### Policy implications

Experiencing well over one million COVID-19 deaths places the United States in a category with Russia, Brazil, and Mexico (20). The real tragedy, however, is the clear regional differences within the country, suggesting that over 400,000 of these deaths could have been avoided. This regional variation, however, also demonstrates the potential impact of population-oriented preventive strategies and other lessons for how the country proceeds in 2022.

The theme of “two Americas” arose in the summer of 2021. At first, the focus was vaccine refusal; later it expanded to opposition to vaccine and mask mandates, and Covid denialism. This pattern was strongest in the South and in states with Republican governors (3,26,27). Using the proportion voting for Donald Trump as a proxy for partisanship, journalistic analyses found consistently higher vaccination rates and lower COVID-19 mortality in Northeastern states, which are predominantly Democratic, and the opposite in Southern states, which are predominantly Republican (4,8,9). But there are other geographic differences in age distribution, racial and ethnic proportions, education levels, and other matters that can be confounding factors (28).

However, rather than starting in the summer of 2021, this analysis demonstrates that large disparities have existed since the beginning of the pandemic. The starkest contrast is between the Northeast and South. In the first use of excess mortality estimates, Woolf and colleagues (15) showed that the first wave of the pandemic was highly concentrated in the Northeast, and particularly in the New York metropolitan area. After the first wave, however, the South, which makes up 38% of the population experienced approximately 45% of all excess deaths. The disparity was most apparent in the summer of 2020, when the daily excess mortality rates were 0.537 per 100,000 in the South and 0.111 per 100,000 in the Northeast. Similarly, the majority (58%) of avoidable COVID-19 deaths between May 31, 2020 and April 30, 2022 were in the South.

Nationally, vaccines have already saved many lives (29) and boosters have the potential to save many more (30). However, our analysis demonstrates that more than half (52%) of the avoidable deaths occurred by the end of February, 2021, when the vaccine rollout was just beginning. Weather is not a likely explanation because the South has had dramatically higher COVID-19 mortality than the Northeast since June, 2020 during all seasons of the year.

Northeasterners may have had higher levels of natural immunity after the first wave (31), but by July, 2020, there were substantially higher seroprevalence rates in the South (37.9%) than in the Northeast (17.5%) (32), so natural immunity cannot explain the large differences starting in the summer of 2020. These results suggest, therefore, that the higher excess COVID-19 mortality in the South and other areas of the country is likely to be due at least in part to higher transmission resulting from differences in mask use, social distancing, and other behaviors. The behavioral survey data in Figure 4 show clear regional differences in the proportion of Americans who wore masks in public all or most of the time during the Alpha wave. The Northeast and West, which had the highest mask utilization rates, had the lowest mortality rates; the opposite was true in the Midwest and South. These differences in mask use remained throughout the study period. The data on individuals working outside the home and avoiding contact with others is less clear cut (see supplemental material). These differences were not attributed to “lockdowns,” which only really characterized the U.S. response in the initial wave. Anecdotal reports suggest strong regional differences whether and when restrictions on public gatherings were lifted (33,34).

**Figure 4.**
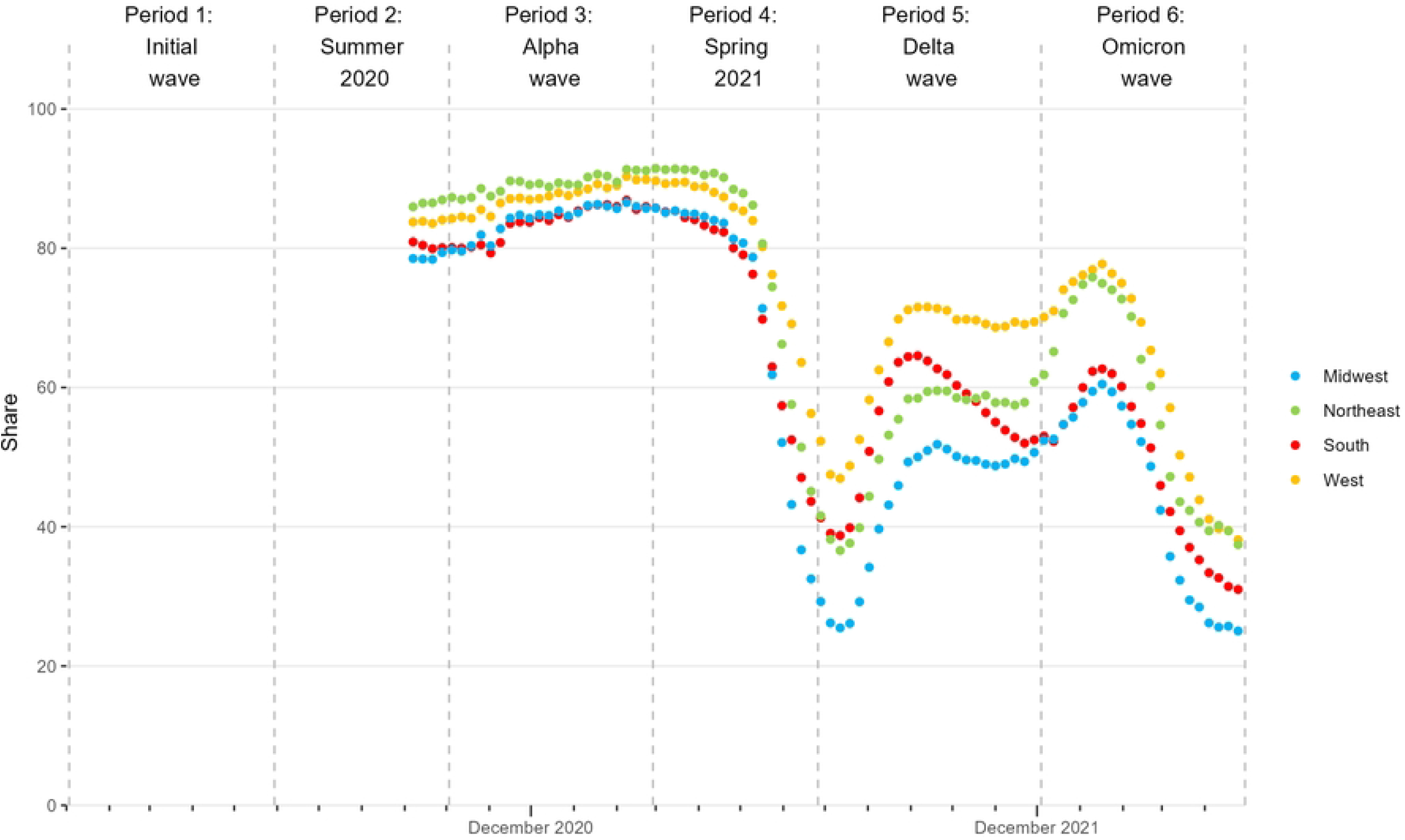
Mask use by period and region, U.S., September 8, 2020 – April 30, 2022. Proportion of survey respondents reporting that they wear a mask in public all or most of the time. Periods defined as in Figure 2. Source: authors’ calculations of Delphi US CTIS survey (see supplemental material).

These behavioral differences could be due to variation in state policies, social attitudes (e.g. regarding “freedom), trust in state and local public health and elected officials, and what these officials said during the pandemic. One analysis found that trust in the federal government was associated with people taking less protective steps (likely because the Trump Administration was undermining control measures during the study period). However, trust in state governments was generally associated with more protective steps, albeit with a complex interaction between individuals’ political affiliation and the party in control of the state government (35).

Regional differences could be partially unrelated to the pandemic. For instance, the Southern states have for many years had higher health risks such as obesity, lower quality health care, and worse health outcomes, and these may have contributed to the observed disparities (16). Sorting out the relative contributions of these factors is beyond the scope of this analysis, which documents the existence of regional differences in COVID-19 mortality and potentially related factors. Calculating excess mortality by state (as opposed to region) as we do in this analysis at least partially controls for persistent differences among states in these factors.

In addition, this analysis translates the correlations between outcomes and potential explanatory factors that dominate both the media and scientific report into concrete numbers of deaths that could have been avoided. Starting in the summer of 2021, strong regional differences emerged in vaccine uptake. Figure 5 shows that, 77% of the population aged 65 and older were fully vaccinated, ranging from 75% in the South to 81% in the Northeast in June 2021. This proportion increased to 90% overall (88% to 94% across regions) in April, 2022. The proportion of those aged 18–64 years who were fully vaccinated was both lower throughout this period, and varied more among the regions. In June 2021 the regional rates ranged from 44% in the South to 61% in the Northeast. Almost a year later (April 2022) the corresponding rates were 67% in the South to 82% in the Northeast. Boosters were not available in June 2021, but in the U.S. the proportion of the fully-vaccinated population aged 65 and older who had at least one booster grew from 41% in December 2021 to April 2022. As with the other vaccine figures, the South lagged behind (37% in December 2021 and 55% in April 2022), but subsequently the Midwest rather than the Northeast had the highest booster coverage rates.

**Figure 5.**
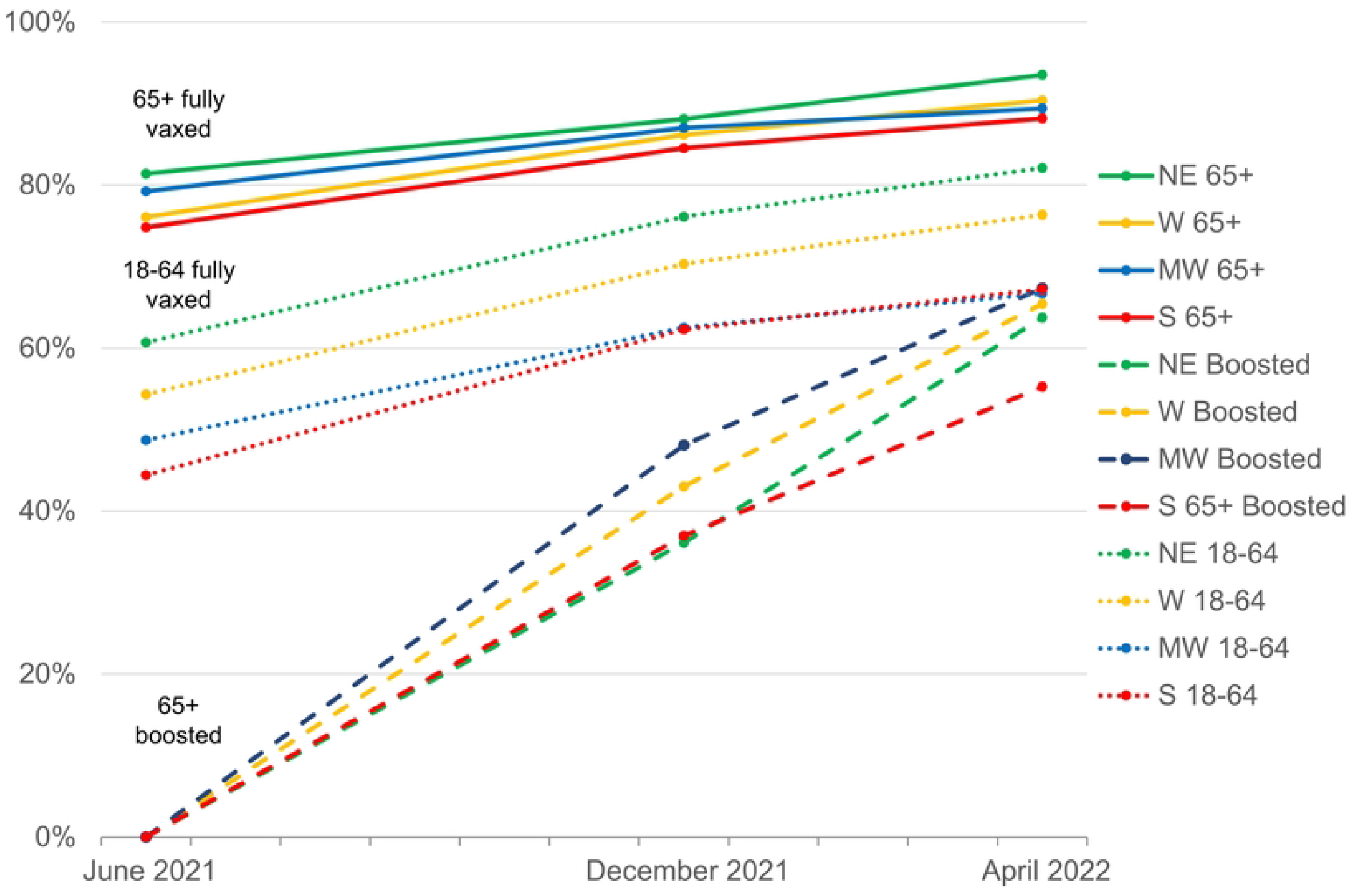
Vaccination uptake by region, U.S., June 26, 2021; December 4, 2021; and April 30, 2022. (1) the proportion of the population aged 65 and older who are fully vaccinated (i.e. have received two doses of the Pfizer-BioNTech or Moderna vaccines or 1 dose of the J&J/Jansen vaccine), (2) the proportion of the population aged 18 to 64 and older who are fully vaccinated, and (3) the proportion of the fully vaccinated population aged 65 and older who have received at least one booster dose. Source: authors’ calculations based on CDC data.

Consequently, regional differences in vaccination likely had a greater impact on mortality during the Delta and Omicron waves (June 27, 2021 – April 30, 2022) than earlier. The vast majority of COVID-19 deaths occur among those aged 65 and older. The proportion of this age group who were fully vaccinated was generally high in all regions, but by late 2021 in the face of the Omicron variant, this protection likely waned, and regional differences in booster rates led to more avoidable deaths, particularly in the South. COVID-19 mortality rates are lower in younger adults, but differences in vaccine coverage rates (from 62% in the South to 76% in the Northeast in December 2021) probably also contributed to differences in avoidable mortality. In addition, although vaccination does not completely prevent infections it does lower the risk (36,37), so differences in vaccine uptake probably added to the impact of non-pharmaceutical interventions in slowing the spread of the virus in some regions. These differences consequently increased the number of high-risk individuals who were infected, hospitalized, and died.

As we write this in the summer of 2022, many are asking when “Covid will be over.” However, with vaccine and booster coverage lagging, especially in rural areas and the regions of the United States that have experienced high COVID-19 mortality, there is still a need for comprehensive, population-based pandemic policies, such as wearing masks in crowded closed spaces. The 2020-style lockdowns are no longer appropriate, but our analysis shows that the less-restrictive measures adopted since the summer of 2020, especially in the Northeast, can still play important roles in protecting those who are most vulnerable to severe disease and death and in reducing the spread of the virus.

### Excess mortality estimates as a component of the surveillance toolbox

Since it emerged in 2020, COVID-19 has played out differently across the country. Beyond the regional differences that are the focus of this paper, there are substantial differences between rural and urban areas with states and among socio-demographic groups (16,38,39). Because they are less sensitive to differences in reporting patterns than case counts, this analysis illustrates how excess mortality methods can be especially useful in understanding these patterns (40). It also shows the importance of going beyond cumulative case counts for the U.S. as a whole that dominate the news cycle.

During the Omicron wave, some observers noted that people were dying “with Covid but not from Covid.” The claim was that testing had become so common that many people who were gravely ill and dying of another cause were included in the COVID-19 deaths counts (41,42). The excess mortality data helps to falsify this claim. While some of the observed mortality may be deaths not directly attributable to COVID-19, the spike in excess deaths beyond what would have been expected in previous years shows that there were substantial numbers of deaths attributable to the pandemic. That observed deaths were concentrated disproportionately among unvaccinated individuals also tends to support this argument: while unvaccinated individuals may take more risks in general, it is hard to imagine that they only drive increasingly recklessly or suffer heart attacks more only when omicron is circulating.

While they cannot explain *why* some regions had different mortality rates, excess mortality estimates are simple and straightforward compared to estimates based on epidemiologic models, and can accurately document *when and where* COVID-19 occurred. In addition, although causal patterns are complex and difficult to ascertain, differential implementation of and adherence to stay-at-home orders, mask use, and other non-pharmaceutical interventions seem to be at least a partial explanation for the regional differences in COVID-19 mortality. Thus, our analysis demonstrates the potential impact of population-based restrictions as well as vaccines, suggesting that a comprehensive COVID-19 policy is needed to control future pandemics.

What this analysis does *not* show is what drives these policy and behavioral changes. For instance, in the summer of 2020, some suggested that state and regional political factors were driving decisions about reopening (43,44). A year later, the same factors drove differences in vaccine uptake (27). Krieger and colleagues (45) suggest that political ideologies of elected representatives drive COVID-19 outcomes. Others have pointed to regional variation in social capital (46,47). Woolf, on the other hand, argues that state mortality differentials during the pandemic simply continue a decades-long trend in political differences between Republican and Democratic dominated states (48) and will continue until the root causes of mortality differentials are addressed (16).

Our analysis also illustrates the importance of analyzing excess mortality (and other COVID-19 data) at the appropriate spatial and temporal scale. With data on cases, hospitalizations, and deaths available on a daily basis for small geographic areas (there are ∼3,000 counties in the U.S.), national patterns can be difficult to see. Add to that multiple and highly variable public health policies and individual behavior changes, assessing the impact of non-pharmaceutical interventions and vaccination campaigns is challenging. Our relatively high-level analysis (four geographical regions and six time periods over two years), has a number of benefits. There have been many studies of the impact of NPIs, but they are limited to a particular context and type of intervention, and by the ability to draw causal conclusions from observational data. Our “big-picture” analysis provides a sense of the scale of NPIs actual and potential impact.

The regional level of analysis we employed strikes a balance between simplicity and objectivity. It is simple enough for disparities to be clearly apparent, but masks more extreme disparities at the state and local levels. For instance, Ackley and colleagues (14) and Paglino and colleagues (49) both found similar regional patterns to this analysis, but additional differences in urban and rural areas. Lundberg and colleagues found racial and ethnic differences in both excess mortality and vaccination acceptance (50). Controlling for demographic characteristics and social determinants likely to influence COVID-19 transmission and outcomes using state fixed effects, Sehgal and others found strong associations between county-level Republican vote share and county-level COVID-19, suggesting that mortality that county-level voting behavior may act as a proxy for compliance with and support of public health measures that would protect residents from COVID-19 (51). These statistics, however, depend on test availability, clinical decisions, and reporting processes that can lead to under– and over– counting. In particular, this analysis shows that the ratio of estimated to reported COVID-19 deaths ranged from 127% (Northeast in period 4) to 52% in the West in Period 5. Thus, the disparities in these analyses are likely underestimates of actual differences at finer levels of geography.

There are, of course, limitations to excess mortality methods. In particular, methods depend on estimates of what mortality rates would have been in the absence of the pandemic. There are many ways of making these estimates, some more appropriate than others (52). Furthermore, a s time goes on, the fundamental assumption that deaths in excess of previous trends are “caused” by the pandemic becomes more tenuous. For example, the death of someone who couldn’t get to an emergency department while experiencing a heart attack in New York City in April 2020 can reasonably be attributed to the pandemic. An opioid overdose in a rural area in April 2022 might be attributed to despair to which the pandemic – and the response to it – contributed, but it is harder to say caused it. As we go further beyond the base period the reliability of these methods will decline.

Excess mortality estimates are limited as real-time surveillance tools. For instance, reporting delays can be greater in some areas than others. Therefore excess mortality estimates and differentials are not reliable for a period of weeks until data are uniformly compiled. On the other hand, hospitalizations and reported COVID-19 deaths lag infections by weeks, and case counts are subject to reporting delays and day-of-the-week effects as well as undercounting. Nevertheless, excess mortality estimates can be a valuable component of a comprehensive surveillance strategy for COVID-19 and other diseases, providing useful information on medium to long term trends – weeks to months.

As some regions cut back on testing because of the sense that the pandemic is “over,” undercounts of deaths — and likely cases and hospitalizations as well — will make it harder to see a re-emergence of COVID-19, particularly BA.5, BA.4.6 and other variants, in those regions. More extensive surveillance based on wastewater testing, seroprevalence surveys, and other means that do not depend on the availability of test sites (53) and individuals’ decisions to be tested is clearly needed to get a more accurate sense of what is happening. Two and a half years into the pandemic, the challenges of predicting expected mortality invalidate excess mortality estimates as real-time surveillance measures, but they can still be useful for retrospective analyses of trends in the pandemic, and the effect of control measures, at the appropriate geographic, socio-demographic, and political scale.

## Data Availability

The excess mortality data, which form the basis of this study, are available from Centers for Disease Control and Prevention: https://www.cdc.gov/nchs/nvss/vsrr/covid19/excess_deaths.htm. Vaccine data are available at https://www.cdc.gov/nchs/data/nvss/vsrg/vsrg03-508.pdf. As noted in the Supplement survey results from Carnegie Mellon University’s Delphi Group. FZ thanks the IRIS Coalition (UK Government, no. SCH-00001-3391). All relevant data for this analysis are in the Supporting Information files.

## Acknowledgments

This research is based in part on survey results from Carnegie Mellon University’s Delphi Group. FZ thanks the IRIS Coalition (UK Government, no. SCH-00001-3391). The authors are grateful to Katrina Dolendo for editorial assistance.

## Notes

### Competing Interest Statement

The authors have declared no competing interest.

### Funding Statement

The authors received no specific funding for this work.

